# A forecasting tool of hospital demand during heat periods: a case study in Bern, Switzerland

**DOI:** 10.1101/2025.11.12.25340087

**Authors:** Laura Di Domenico, Martin S. Wohlfender, Wolf E. Hautz, Ana M. Vicedo-Cabrera, Christian L. Althaus

**Affiliations:** Institute of Social and Preventive Medicine, University of Bern, Bern, Switzerland; Multidisciplinary Center for Infectious Diseases, University of Bern, Bern, Switzerland; Department of Emergency Medicine, Inselspital University Hospital Bern, Bern, Switzerland; Oeschger Center for Climate Change Research, University of Bern, Bern, Switzerland; Graduate School for Cellular and Biomedical Sciences, University of Bern, Bern, Switzerland

## Abstract

**Introduction:** Heat significantly impacts human health by causing heat strain or exacerbating pre-existing conditions. Hospitals may suffer a higher healthcare demand during intense heat periods, especially if climate change continues to increase the severity and frequency of heatwaves. Anticipating episodes of higher hospital demand would allow better resource planning and quality of care.

**Methods:** We developed a real-time forecasting tool of daily hospital demand (specifically, all-cause emergency room visits (ERV)) which accounts for the impact of heat. Our tool is based on a regression model integrating temperature-ERV function with autoregressive terms and other temporal trends. The model can (i) quantify the association between the number of hospital visits and temperature based on historical data and (ii) provide accurate short-term forecasts of the daily ERV based on temperature values expected for the upcoming days. As a case study, we used data from the Bern University Hospital from the summer of 2014 to 2022, and mean temperature per day as an indicator of heat exposure.

**Results:** Temperature-ERV relationship exhibited a non-linear shape. We found that, with respect to the mean temperature of minimum risk of 15°C, there were approximately 6 (95% CI 2-10) additional ERVs when mean temperature was around 25 °C, corresponding to a 3% increase in summer 2022. The estimated variation increased for mean temperature above 25 °C, but with large uncertainty. We also found that our model showed higher accuracy at forecasting hospital demand during periods with particularly hot days, compared to a model neglecting temperature. Our forecasting tool is implemented in a user-friendly R shiny app, allowing for application to new datasets.

**Conclusions:** We found a robust association between ambient temperature and visits to the emergency department in a Swiss hospital. Our findings suggest that including temperature can increase the accuracy of predictions for hospital demand during summer.

## Introduction

Exposure to high temperatures significantly impacts human health, causing heat-related physiological stress or exacerbating pre-existing conditions such as cardiovascular diseases and respiratory disorders like asthma^1^. Extreme heat episodes have been increasingly recognized as a serious public health threat, especially in the context of climate change with frequency and severity of heatwaves expected to increase in the future^2^. Furthermore, the increasing age of many populations aggravates the problem as older people are substantially less heat tolerant than younger ones^3^. However, the potential impact of climate change on healthcare workers and the quality of care remains unclear. Numerous studies quantified the association between high temperature and increased morbidity, including hospitalizations and visits to the emergency room^4–7^. Therefore, healthcare infrastructure may suffer under future scenarios of high warming with periods of very intense heat. This has become even more relevant, since recent public health crises such as the COVID-19 pandemic showed how vulnerable the healthcare system can be in periods of extremely high demand. Thus, forecasts of hospital demand during heat periods can be useful for resource planning, especially for emergency departments to avoid the ubiquitous problem of overcrowding and reduced quality of care^8,9^.

By using standard methods in health impact assessment, recent epidemiological investigations estimated the mortality and morbidity burden due to heat in different study settings^10,11^. However, very few studies exploited this relationship to produce short-term forecasts^12^. In this study, we developed a forecasting tool of the number of hospital visits during summer, accounting for the impact of heat. Our tool is based on a regression model that combines time-series predictors (e.g. autoregressive terms, weekly pattern) and daily temperature through a distributed lag non-linear model. As a case study, we applied our forecasting model to predict the number of emergency room visits (ERVs) during summer in Bern University Hospital, one of five first-level university general hospitals of Switzerland.

We obtained the exposure-response function empirically using observed weather and health data, and assessed the accuracy of short-term forecasts of hospital demand, both including and omitting temperature in the model. We implemented our tool in a user-friendly R shiny app, that can be easily used and extended in other case studies.

## Materials and Methods

### Data sources

#### Hospital data

We used routinely collected individual-level electronic health records from the Bern University Hospital in the city of Bern, Switzerland. We extracted the daily number of patients visiting the emergency department, for any cause (all-cause emergency-room visits, ERV). The number includes both ambulatory patients (discharged within the day) and patients admitted to the hospital for at least one night. We considered historical data for the period ranging from 2014 to 2022. We restricted our analysis to the warmest months of the year, defined from June 1st to August 31st.

#### Temperature data

We used historical data of daily mean temperature, registered at the Zollikofen weather station between 2014 and 2022, the closest station to the city of Bern. Data was requested through the IDAweb portal^13^, maintained by Meteo Swiss. To evaluate forecasts, we first used the observed temperature on the test set. To simulate a real-time application, we then used forecasted temperature data provided by Meteo Swiss. To ensure comparison of the temperature grid data with the observed historical data, we considered a grid point in the surroundings of the Zollikofen measurement station and bias-corrected for altitude difference. Besides mean temperature per day, we also considered other temperature indicators (i.e. daily and nocturnal means and minimum and maximum temperature) in a sensitivity analysis (Fig. S5).

### Statistical analysis

#### Statistical model

Our model included both autoregressive terms and explanatory variables as model predictors^14^. The outcome variable was the daily number of ERV. In short, the model included the following variables: an intercept, previous ERV up to seven days in the past, day of the week and holiday effect, a level shift in the 2020-2022 period to account for COVID-19 effect, and lagged temperature values from lag 0 to lag 3. This allows the model to capture short-term effects of temperature and harvesting effect. For temperature, we used an unconstrained distributed lag nonlinear model^15^, meaning that each lagged temperature was modeled with a non-linear function (natural splines). The full mathematical description of the model is presented in the Supplementary Information. We then computed the cumulative exposure-response function by aggregating over the lags. We did not put constraints in the lag-response dimension, meaning that each lagged temperature was modeled independently. This choice was adopted to ease implementation for forecasting purposes.

We fitted the regression model assuming a Gaussian distribution. We found that the shape of the exposure-response function estimated from our dataset was not sensitive to the choice of the family (Gaussian or quasi-Poisson, Fig. S6).

#### Model calibration and evaluation

We estimated the exposure-response function linking ERV counts and temperature by training the model on data from 2014 to 2021. We used data from summer 2022 as a test set. We also carried out sensitivity analysis with cross validation (i.e., using a different year as test set, and the remaining years as training set) to check the robustness of the shape of the exposure-response curve (Fig. S4). To evaluate model accuracy, we computed the root mean square error (RMSE) between the 1-step ahead model prediction 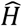(𝑡) and the observed data 𝐻(𝑡) on day 𝑡. We computed the average RMSE on the training set and on the test set. Model accuracy was then compared with a model without temperature, and basic LOCF (last-observation-carried-forward) models with a 7 day lag or with a 1 day lag (i.e., the prediction 𝐻(𝑡) for day 𝑡 is equal to 𝐻(𝑡 − 7) or 𝐻(𝑡 − 1), respectively).

#### Forecasts

We generated ℎ-step ahead forecasts, for forecast windows of one, two or three weeks ahead. We generated point predictions and prediction intervals (PI) at 50% and 90%. We evaluated forecast errors of the point predictions using average RMSE over the forecast window, and compared them with results obtained using a model without temperature.

Comparison of the two models was also framed as a function of the average temperature in the forecast window, to determine which model performed better specifically over forecast windows with a high average temperature. We generated the forecasts retrospectively on the test set (summer 2022), using available historical data for temperature. To simulate a real-time use of the forecasting tool, we used forecasted temperature data for summer 2022, and compared model outcomes with the results obtained using observed temperature data.

## Results

### Estimation of the association of ERV with temperature

We used historical data on emergency room visits (ERV) and mean temperature per day from 2014 to 2022 (Fig. 1). The time-series of ERV showed a significant level shift in the period ranging from 2020 to 2022, during the COVID-19 pandemic (Fig. 1a,b). The periods registering the largest number of days with particular high temperature (>25 °C) were summer 2015 and 2019 (Fig. 1c,d, Fig. S1).

**Figure 1.**
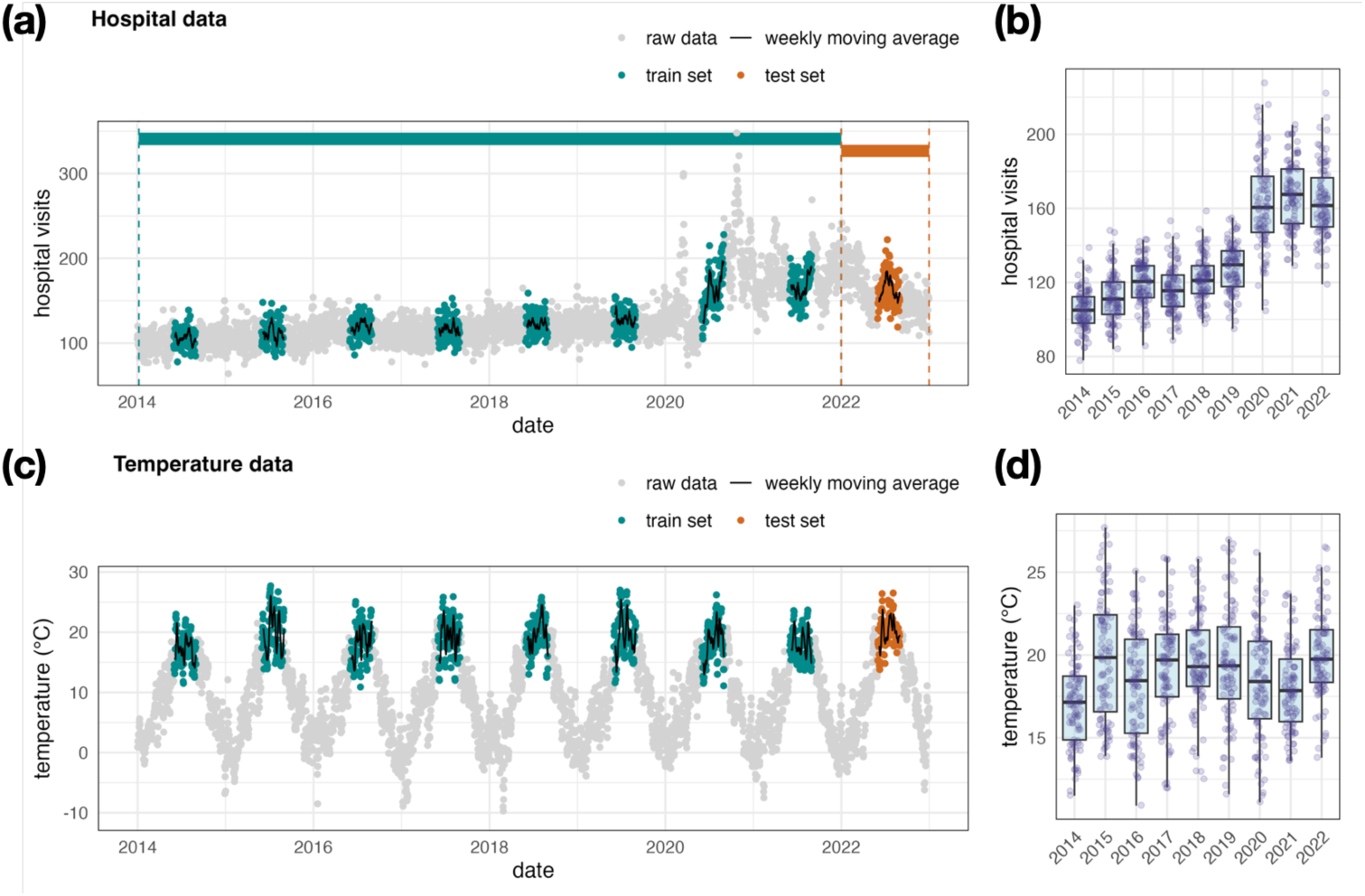
Data. **(a)** Time-series of daily emergency room visits (ERV); **(b)** Boxplot (median and interquartile range) showing the distribution per year. **(c)** Time-series of the observed mean temperature per day, expressed in °C. **(d)** Boxplot showing the distribution per year. In panels (a) and (c), grey dots represent the full time-series, colored dots represent summer data used for the analysis (from June 1st to August 31st). Green and red datasets indicate the training set and the test set, respectively.

We fitted our model to a training set ranging from 2014 to 2021. Model predictions (1-step ahead) for each year of the training set are shown in Fig. S3 in the Supplementary Information. We found that the Akaike Information Criterion (AIC) for the model including temperature was slightly lower compared to the model neglecting temperature (Table S1). We computed the root-mean-squared-error (RMSE) between the 1-step ahead model prediction and the observed data on the training set and the test set (summer 2022). We found that the error of our model was much lower than the error of last-observation-carried-forward models. The error was similar to a model with the same set of predictors but removing temperature. These results are summarised in Table S2.

We computed the cumulative exposure-response function (Fig. 2a), measuring the non-linear association between ERV and temperature, by aggregating the effects of lags up to three days in the past (Fig. 2b). We found that, compared to a mean temperature of minimum risk of 15 °C (as estimated by the model), there were approximately 6 (95% CI 2-10) additional ERV when mean temperature was between 20-25 °C. This number corresponded to a 3% (95% CI 1-6%) relative increase over the average number of visits registered during summer 2022 (during COVID-19 period), or 5% (95% CI 2-9%) over the average observed before COVID-19. The estimated median variation increased for extremely high temperatures (28 °C), although with large uncertainty due to the low occurrence of such values in the training set (Fig. 2c). We found that the shape of the exposure-response function was stable after cross-validation on the training set (Fig. S4).

**Figure 2.**
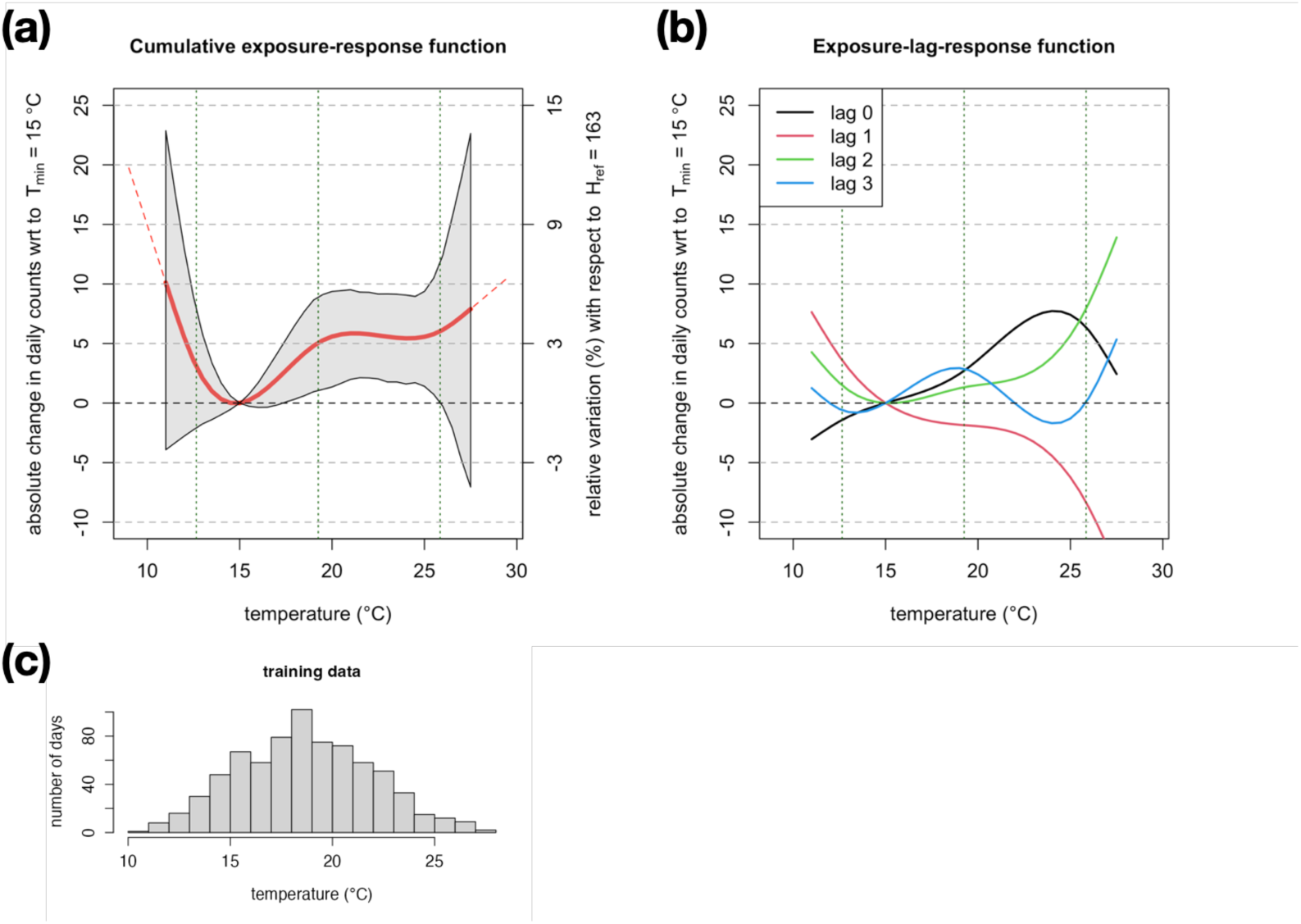
Estimated non-linear association with mean temperature per day. **(a)** Cumulative exposure-response function, showing the absolute change in daily hospital visits as a function of temperature, with respect to a reference temperature of minimum risk of 15 °C. On the right y-axis, relative variation with respect to the average number of visits registered during summer 2022 (test set). **(b)** Lag-exposure-response function for different lags. In panels (a) and (b), vertical dashed green lines indicate the position of the internal knots used to define the non-linear spline functions. **(c)** Histogram of the temperature values observed in the training set.

### Forecasts

We used our model to produce forecasts of daily ERV up to 3 weeks ahead for the testing period (2022), and retrospectively evaluated forecast errors using the observed data as reference (Fig. 3). The model is able to capture changes in trends of patient visits (Fig. 3a). The model is more precise in predicting the pattern of hospital visits in the upcoming week, while errors become increasingly larger when evaluating predictions two or three weeks in the future (Fig. 3b).

**Figure 3.**
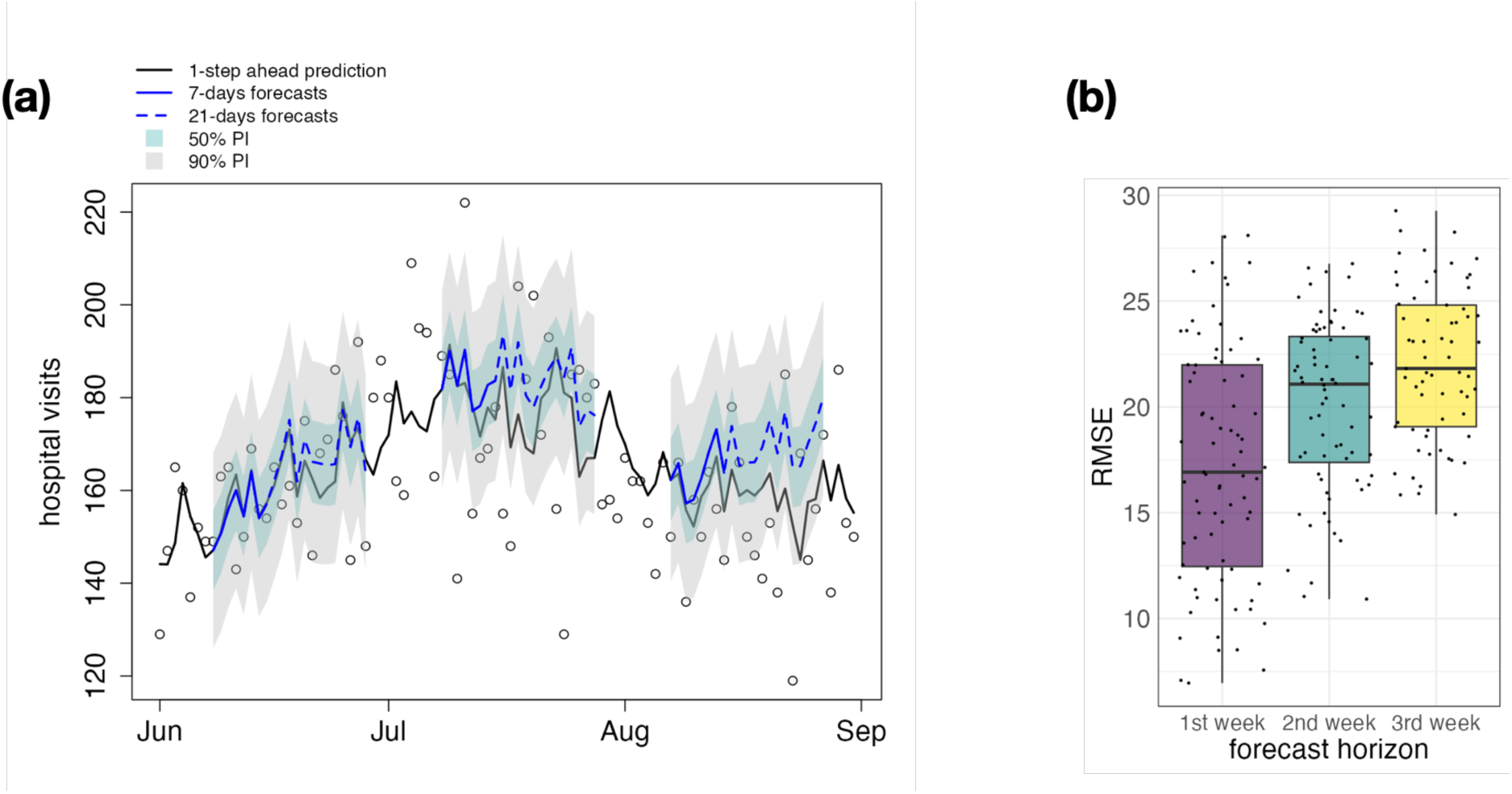
Forecasts of daily ERV with different time horizons. Example using summer 2022 as test set. For each given forecasting horizon (1, 2 or 3 weeks), we consider time-windows with different starting dates, spanning from June 8th up to the end of August. **(a)** Daily number of hospital visits (specifically, emergency-room visits). Three forecast windows (with starting date in June, July or August) are shown as examples. Blue lines show the estimated model prediction, along with 50% and 90% prediction intervals (PI) shown with shaded areas. In black, we show the 1-step ahead model prediction. **(b)** Forecast error evaluated over a period of 7 days, in the first, second or third week from the starting date. The boxplot indicates the median and the interquartiles.

We also generated forecasts of daily ERV using forecasted temperature data, to simulate the use of the tool in real time. We found that the forecast errors remained similar when replacing observed temperature data with forecasted temperature data (Fig. S10), provided that the two sources are in good agreement (Fig. S8). We also found that integrating the uncertainty in the forecasted temperature data (with variations around 2 °C, Fig. S7) in the hospital forecasting model did not significantly increase the width of the prediction intervals (Fig. S9).

Finally, we aimed to assess whether the proposed model provided more accurate forecasts compared to a model neglecting temperature. To increase the sample sizes, we generated forecasts ranging from 2014 to 2022, using models trained on all data except the year of the forecasts (i.e., using cross-validation on the training set). We compared the RMSE of the forecasts with respect to a model neglecting temperature. First, we considered all forecast windows. We found that the median RMSE was similar for the first week of prediction, and slightly smaller for the second or third week of predictions with respect to a model neglecting temperature (Fig. 4a). Then, by selecting only the forecast windows for which the average temperature was quite high, we found that the median RMSE obtained with the proposed model was lower than a model without temperature (Fig. 4b). This is illustrated in Fig. 4b using a threshold of 22 °C, but this trend was observed for any temperature threshold larger than 20 °C. Overall, these results suggest that, although for middle-low temperature values the two models perform similarly, a model including temperature is more accurate when forecasting over periods with particularly hot days compared to a model neglecting temperature.

**Figure 4.**
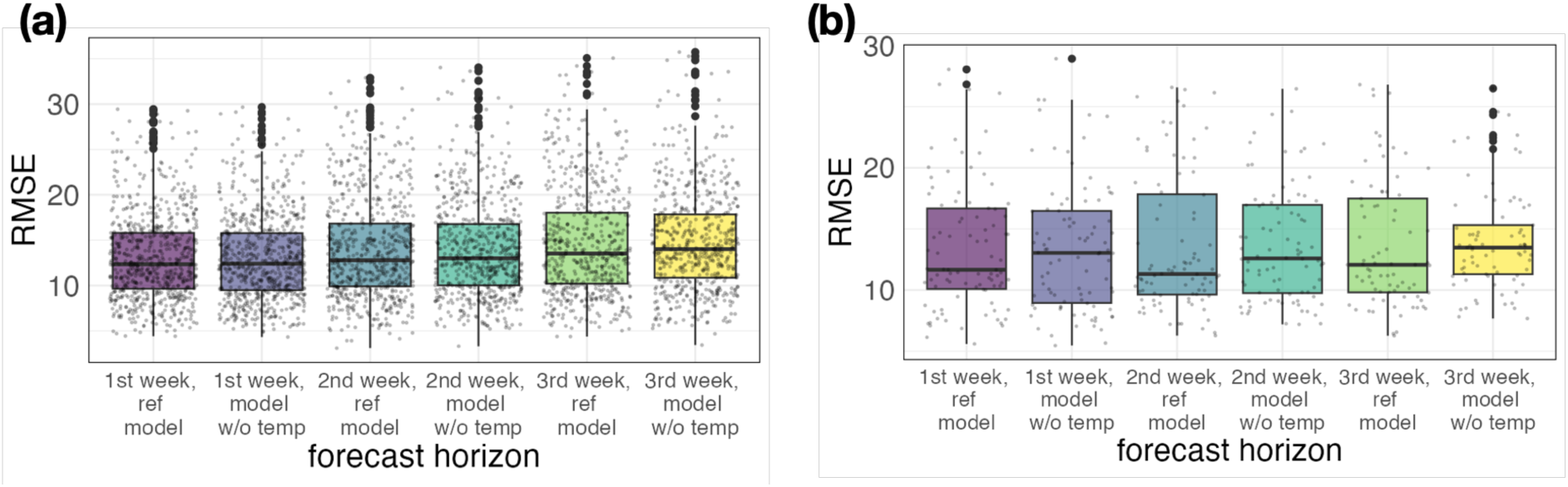
Comparison of forecast errors for a model with or without temperature. For each given forecasting horizon (1, 2 or 3 weeks), we consider 65 time-windows, with the starting date spanning from June 8th to August 11th. We generate forecasts for each year (from 2014 to 2022), excluding the year of the test set from the training data. **(a)** All forecast windows. **(b)** Only forecast windows with average temperature above 22 in °C.

## Discussion

Here we present a method and tool to predict future hospital demand in terms of ERV accounting for the effect of temperature. We estimated the non-linear relationship between temperature and hospital visits, using state-of-the-art methods and historical data, and leveraged this relationship to generate forecasts of expected hospital demand for resource planning. To achieve this, we adopted a novel approach that combines a distributed lag non-linear model with a time-series forecasting framework. In our case study, we found a non-negligible association between observed temperature and ERVs in a major hospital in Bern, Switzerland. Temperature above 20 °C corresponded to an increase of at least 3% in the daily ERVs compared to an optimal temperature of reference (estimated at 15 °C). The model was able to reproduce historical trends in ERVs. Short-term forecasts (1-3 weeks ahead) showed higher accuracy over forecast horizons with elevated average temperature, compared to a model neglecting temperature, suggesting that our model more effectively captured the impact of heatwaves on hospital demand.

We implemented our model as a user-friendly R Shiny application, facilitating its application to other case studies, and enabling potential real-time use when informed with forecasted temperature data. While we demonstrated our approach using emergency department data, the model is easily adaptable to other datasets, e.g. visits to other hospital wards or patients with specific diagnoses. Additionally, our tool allows the user to estimate the corresponding location-specific relationship between hospital visits and temperature using observed data, which is crucial given that this relationship may vary across settings. Forecasts are then generated using this specific exposure-response function. Finally, our framework allows for testing various temperature indicators. Although mean daily temperature was used in our main analysis, alternative metrics may also be relevant, e.g. nocturnal heat which has been found to be associated with increased risks of stroke^16^.

Our study has some limitations. First, the model was trained on a limited number of years, and heatwave episodes were quite rare; more data encompassing extreme heat periods would allow to refine the model estimates. A key assumption of our forecasting tool relates to the exposure-response function used to generate forecasts. This function is estimated on historical data and is assumed to be representative of current and future heat-related risks, meaning that our framework does not account for adaptation to heat or increased heat intolerance in an aging population. Incorporating an exposure-response function which evolves over time could help account for potential adaptation mechanisms, but as of today, the required direction and extent of such a model adaptation remains unknown. Another limitation concerns the impact of the COVID-19 pandemic. We assumed that, from 2022 onwards, the average hospital demand returned to pre-pandemic levels (prior to 2020). This assumption was met for our case study but may not be optimal for all datasets. Forecast errors were assessed using only the point estimates, without considering the full range of uncertainty; future studies could adopt evaluation metrics that explicitly incorporate predictive uncertainty. Finally, while we integrated forecasted temperature data into our model, we did not retrospectively assess the association between ERVs and forecasted temperature values. This was not feasible for our case study, due to the availability of only one summer of forecasted temperature data. Nevertheless, the comparison of forecasted and observed temperature data indicated good agreement, suggesting that bias correction was not necessary.

Our study identifies heat as a relevant driver for emergency department visits during summer, and suggests that inclusion of temperature can increase the accuracy of forecasts of hospital demand during heatwaves. Forecasting tools that account for this effect would be particularly relevant for hospital capacity planning, especially in the context of current projections of climate change, with more frequent and intense heatwaves expected in the future. The forecasting tool for emergency department visits developed in this study offers a foundation for broader implementation in real-time settings and future integration in the management infrastructure of healthcare facilities.

### Data and code availability

The code of the forecasting tool, together with the daily time-series of ERV and temperature, are publicly available on GitHub at https://github.com/ISPMBern/hospital_forecasting_tool (Ref.^17^). The web-application is accessible at https://hospital-forecasting-tool.ispm.unibe.ch/.

### Author contributions

A.M.V.C. and C.L.A. conceived the study. L.D.D. and C.L.A. developed the methodology. L.D.D. developed the software for the forecasting tool, performed the statistical analysis, and analysed the results. M.S.W. tested and validated the forecasting tool. L.D.D. wrote the first draft of the manuscript. All authors critically revised the manuscript and approved its final version.

### Competing interests

The authors declare no competing interests.

## Data Availability

The code of the forecasting tool, together with data on the daily time-series of ERV and temperature, are publicly available on GitHub.

https://github.com/ISPMBern/hospital_forecasting_tool

## Acknowledgements

We acknowledge Evelyn Mühlhofer (MeteoSwiss) for providing forecasted temperature data, and Marcos Quijal Zamorano (University of Bern) for testing the forecasting tool. We gratefully acknowledge the Insel Data Science Center (IDSC) (www.idsc.io/en/) for facilitating the access to the electronic health records from the Insel Gruppe hospital network.

## Funding Statement

This study received funding from the National Centre of Climate Services through the NCCS-Impacts program, project “Impact of climate change on human and animal health”. The work was further supported by the Multidisciplinary Center for Infectious Diseases, University of Bern, Bern, Switzerland. A.M.V.C. acknowledges funding from the Swiss National Science Foundation (TMSGI3_211626) and from Mobiliar Cooperative. The latter had no role in the conception and development of this work.

## Ethical approval

Ethical approval of the study was granted by the Health, Social and Integration Directorate Cantonal Ethics Committee for Research of the canton of Bern (number 2022-01692).

## Supplementary Information

### Temperature data

The seasons registering the largest number of days with mean temperature above 25 °C were summer 2015 and summer 2019, with 9 and 6 days respectively. For context, the 25 °C threshold is currently being used by MeteoSwiss to issue heat warnings in Switzerland^18^.

**Figure S1.**
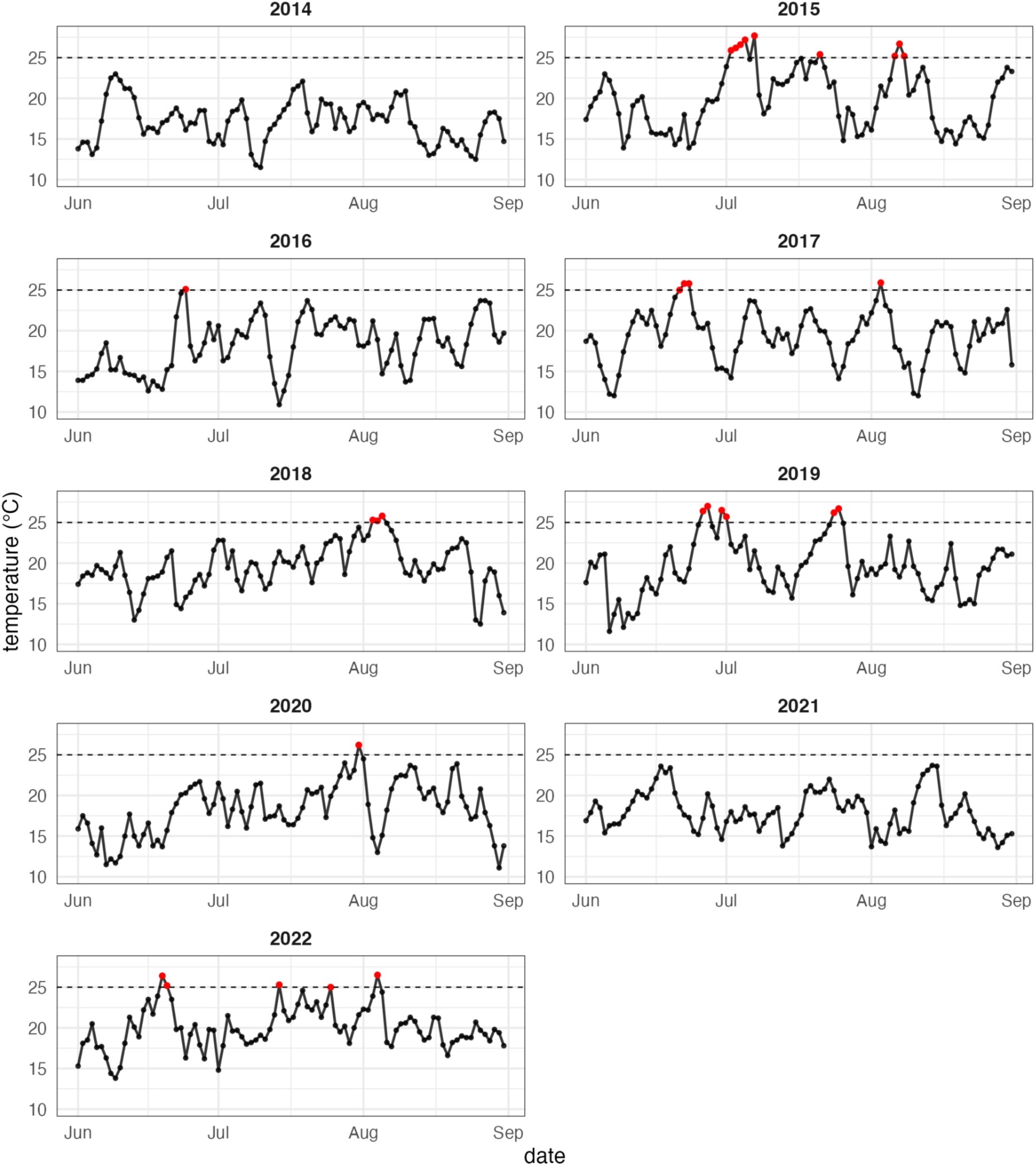
Temperature data. Each plot shows the daily time-series of mean temperature, for each year ranging from 2014 to 2022, from June 1st to August 31st. Days with temperature values exceeding 25 °C are highlighted in red.

### Model description

The model can be summarised as follows

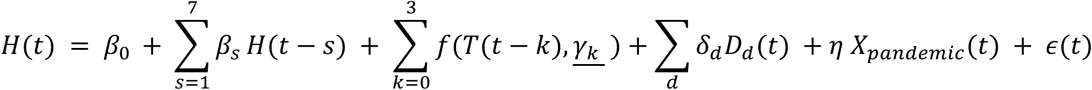

where 𝐻(𝑡) is the outcome, i.e. the daily number of ERV on day 𝑡, and 𝜖(𝑡) represent the error terms. The coefficient 𝛽_!_ denotes the intercept. The autoregressive terms 𝐻(𝑡 − 𝑠) refer to daily observations of hospital visits up to seven days in the past. To model the association with temperature 𝑇(𝑡), we used an unconstrained distributed lag non-linear model^15^. In detail, we used a natural spline function 𝑓 (i.e., a cubic spline with constraints of linearity beyond the boundary knots^19^), for each 𝑇(𝑡 − 𝑘) from current day 𝑡 up to 𝑘 = 3 days in the past. This allows the model to capture short-term effects of temperature and harvesting effect. For each lag, we used three internal knots for the spline function, i.e the median and 10% and 90% quantiles of the observed temperature range, for a total of four degrees of freedom. Hence, we estimated four parameters 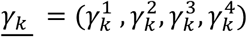 for each lag. We then computed the cumulative exposure-response function by aggregating over the lags. We did not put constraints in the lag-response dimension, meaning that each lagged temperature was modeled independently. This choice was adopted to ease implementation for forecasting purposes. Finally, we included the following control variables for temporal trends. We included seven dummy variables 𝐷*_d_*(𝑡) ∈ {0,1} to account for the day of the week, distinguishing between regular weekdays (𝑑 = Tuesday, …, Friday), Saturday, Sunday and holiday within the week; Monday was used as reference. We included a dummy variable 𝑋*_pandemic_*(𝑡) which equals to 1 in the period from 2020 to 2022, to capture a level shift during the COVID-19 pandemic. The inclusion of these temporal variables was based on improvement of the Akaike Information criterion (AIC) (Table S1). For sensitivity, we also tested the inclusion of a within-year monthly seasonality, but this did not improve the AIC.

To generate ℎ-step ahead forecasts retrospectively on the test set, we adopted the following procedure. Let day 𝑡 be the start of our forecast window, and let 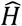(𝑡) be the 1-step ahead prediction obtained with the model described above. To extend our forecasts, we used 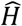(𝑡) as autoregressive term to compute 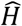(𝑡 + 1), using the observed temperature data *T*(𝑡 +1) as input, and so on up to 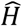(𝑡 + ℎ − 1). To compute forecasts in real time starting from day 𝑡, forecasted temperature data 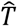 (𝑡 + 𝑘) for 𝑘 = 0, . . ., ℎ − 1 are needed to feed the model.

### Model selection

We compared our proposed model with variations of the model removing some predictors (temperature, weekly pattern, COVID-19 shift) or including additional temporal effects (month seasonality). In this set of models, our proposed model showed the best goodness of fit (lowest AIC). There was strong evidence against model without weekly pattern and COVID-19 level shift (𝛥𝐴𝐼𝐶 > 20), while the other models scored with a 𝛥𝐴𝐼𝐶 around 2 or 3.

**Table S1.** Akaike Information Criterion (AIC) for different models.

| Model | AIC |
| --- | --- |
| Proposed model | 5858.0 |
| Proposed model without temperature | 5860.8 |
| Proposed model without day of the week & holiday effect | 5953.9 |
| Proposed model without level shift due to COVID-19 | 5880.7 |
| Proposed model with month indicator | 5859.9 |

### Model coefficients, fit and evaluation

Besides the estimated coefficients related to temperature, we can also extract coefficients related to other predictor variables, in order to provide some interpretability of the model (Fig. S2). Our model included seven autoregressive terms, i.e. the daily number of hospital visits observed in the last seven days. We found that the corresponding coefficients decrease the longer the lag. This suggests that daily counts observed in less recent days play a smaller role in predictions with respect to more recent days. Our model also included a dummy variable for the day of the week, distinguishing between regular weekdays (from Monday to Friday excluding holidays), weekends (Saturday and Sunday) and holiday. We found that, using Monday as a reference, there are fewer hospital visits on Tuesday, and the number gradually increases during the week until Saturday. The number of expected visits on a Saturday is comparable to a Monday. Fewer hospital visits are associated with Sunday and bank holidays.

**Figure S2.**
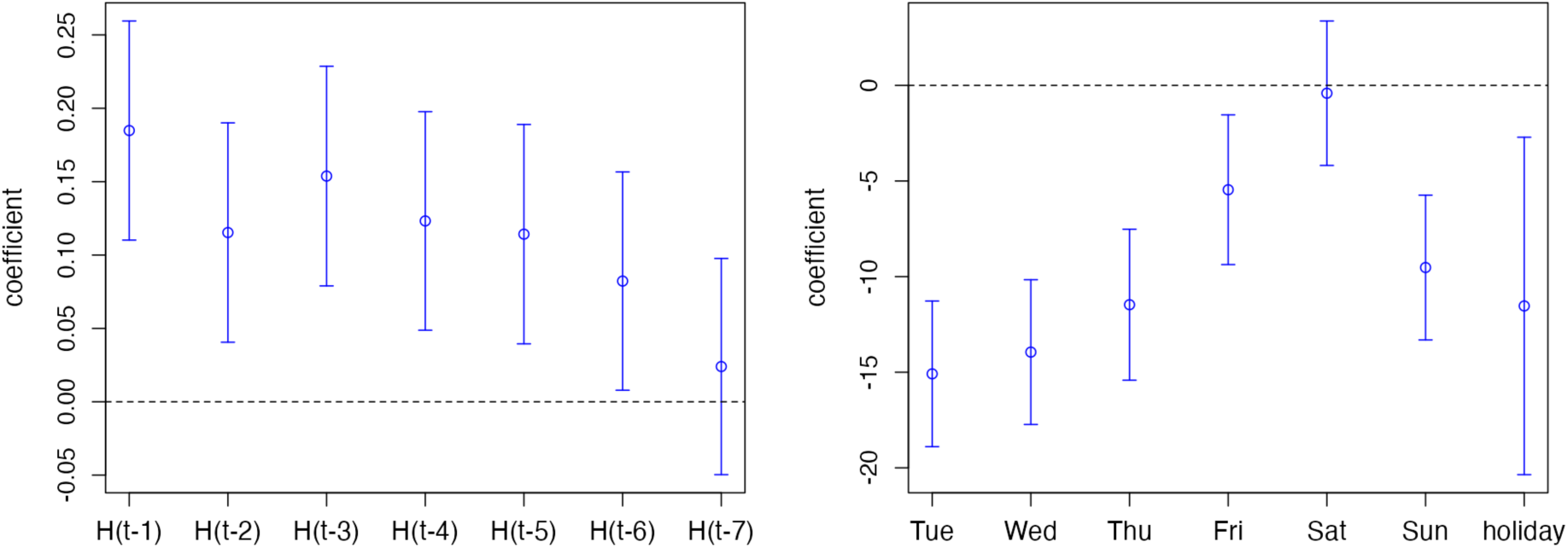
Left panel: estimated coefficients for the autoregressive terms (defined in the Methods as 𝛽*_s_* with 𝑠 = 1, . . .,7). Right panel: estimated coefficients for the dummy variables describing the weekly pattern (defined in the Methods as as 𝛿*_d_*); Monday is used as a reference.

In Fig. S3, we show the results of the model fitted to the training data. The 1-step ahead model predictions are able to follow the trends in the data, with a short delay.

**Figure S3.**
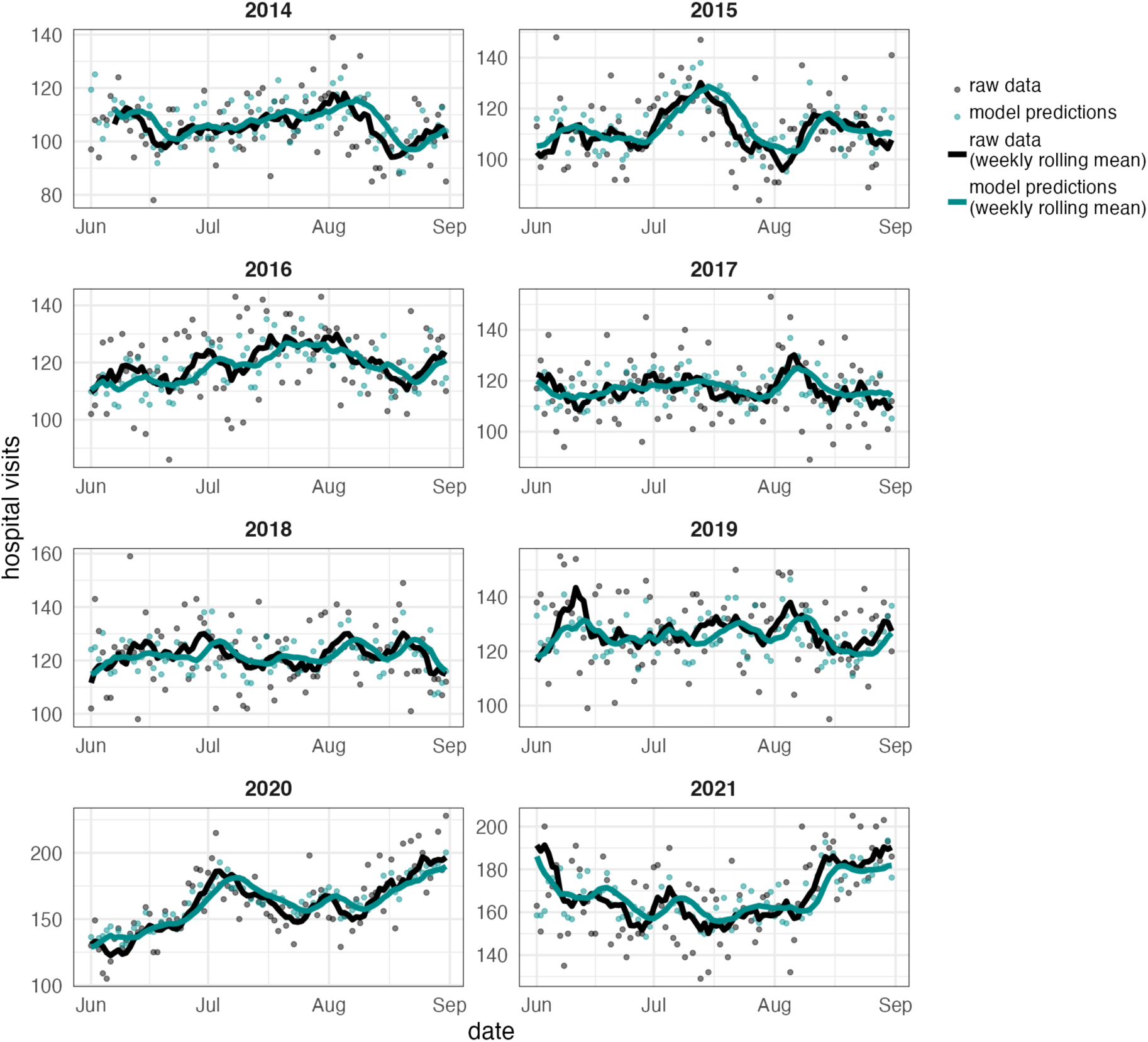
Model fit on the training set. For each year in the training set, we show the daily observations (black dots) and model estimates (1-step ahead prediction, green dots). Weekly rolling mean for both time-series is shown with a continuous line, as a guide for the eye.

In Table S2, we show the root-mean-squared error computed between the 1-step ahead point prediction of the model and the observations. We computed this metric on both the training and the test set. We compared the results of our model with a model neglecting temperature and two last-observation-carried-forward (LOCF) models, with 1-day or 7-day lag. We found that the error of our model was much lower than both LOCF models. The error was similar compared to a model with the same set of predictors but removing temperature. To make a fair comparison of the RMSE across train set (2014-2021) and test set (2022, during the COVID-19 pandemic), we distinguished between the subset of training data which refer to the pre-pandemic period (prior to 2020) and the period of the COVID-19 pandemic (2020-2021).

**Table S2.** Root-mean-squared-error (RMSE) between the 1-step ahead point predictions of different models and the daily observations. The RMSE is stratified by training and test set, and further stratified distinguishing the training set prior or during the COVID-19 pandemic.

|  | RMSE |  |  |  |
| --- | --- | --- | --- | --- |
| Model | Train set<br>(2014-2021) | Train set<br>(2014-2019) | Train set<br>(2020-2021,<br>COVID-19) | Test set<br>(2022, COVID-19) |
| Proposed model | 12.37 | 11.21 | 15.35 | 17.36 |
| Proposed model<br>without temperature | 12.67 | 11.48 | 15.71 | 16.87 |
| LOCF (7 day lag) | 17.82 | 16.00 | 22.43 | 18.09 |
| LOCF (1 day lag) | 17.60 | 16.53 | 20.48 | 25.75 |

### Exposure-response function with cross-validation

In Fig. 2 in the main text, we showed the exposure-response function estimated using data from 2014 to 2021 as a training set, excluding data for 2022. In this section, we present the estimated exposure-response function resulting from the inclusion of all available years, or exclusion of one single year other than 2022. This cross-validation analysis shows that the shape of the exposure-response function is fairly robust. The major differences are found in the estimate of the right-end tail (mean temperature above 25 °C) when excluding data from 2019 or 2015 from the training set. This is due to the fact that these years correspond to the seasons who registered the highest number of days with high temperature (Fig. S1), hence removing these data from the training sets has an impact on the estimates of the right-end tail of the exposure-response function.

**Figure S4.**
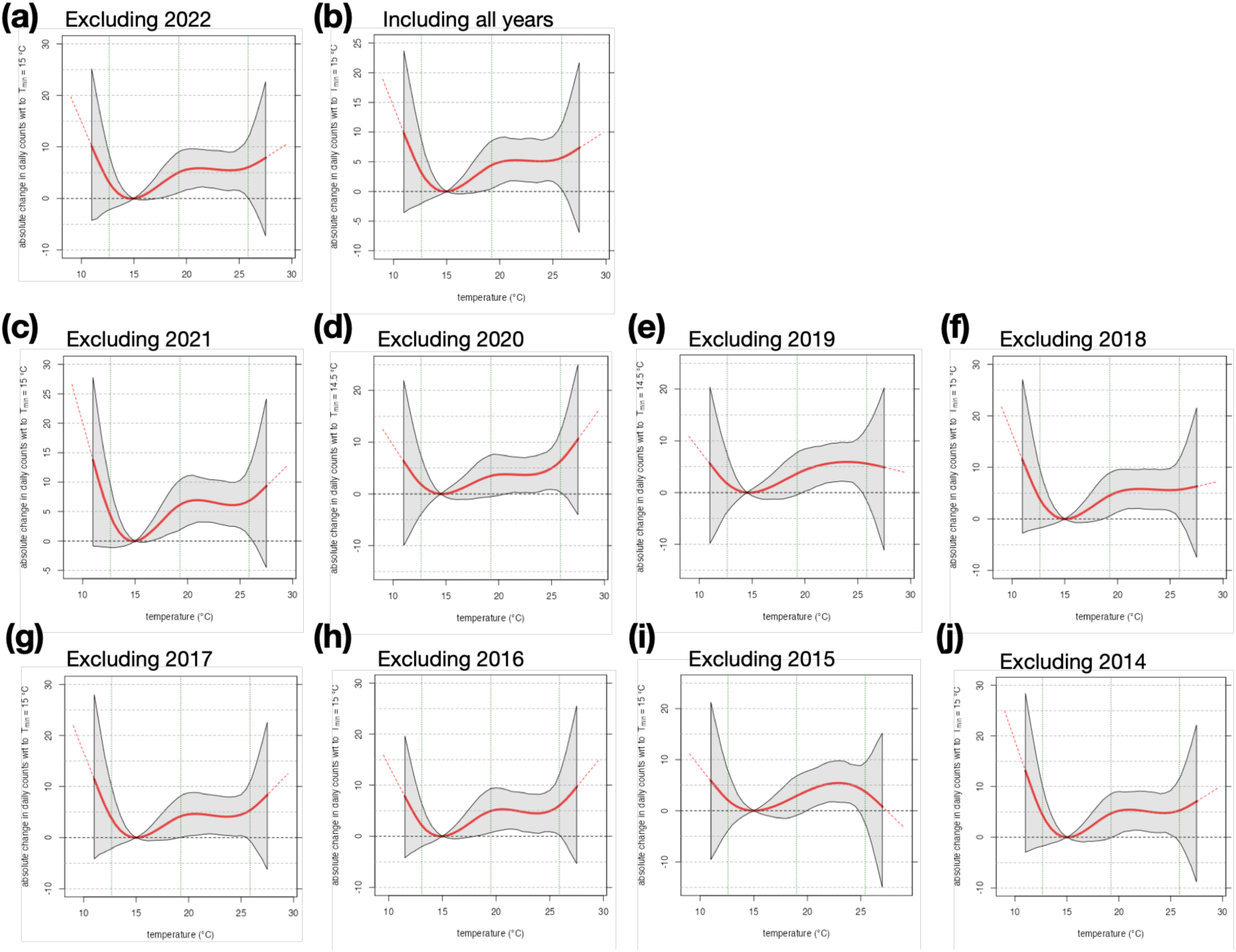
Estimated non-linear association with temperature, after cross validation on the training set. **(a)** Cumulative exposure-response function presented in the main text (Fig. 2a) obtained excluding data for 2022 from the training set. **(b)** Cumulative exposure-response function obtained using all available data (2014-2022) as training set. **(c-j)** Cumulative exposure-response function obtained using all data except one year (either 2021, 2020, …, or 2014).

### Other temperature indicators as exposure variable

In this section, we provide the results of the estimated association between hospital visits and temperature, using different temperature indicators. In the main text, we used mean temperature per day. Notably, we found that replacing the exposure indicator with mean temperature during the day (6am - 6pm) or during the night (6pm - 6am), or maximum temperature, led to similar results in terms of the shape of the estimated cumulative exposure-response function. In contrast, using minimum temperature as the exposure indicator led to highly uncertain results, suggesting that this indicator does not capture relevant heat effects.

**Figure S5.**
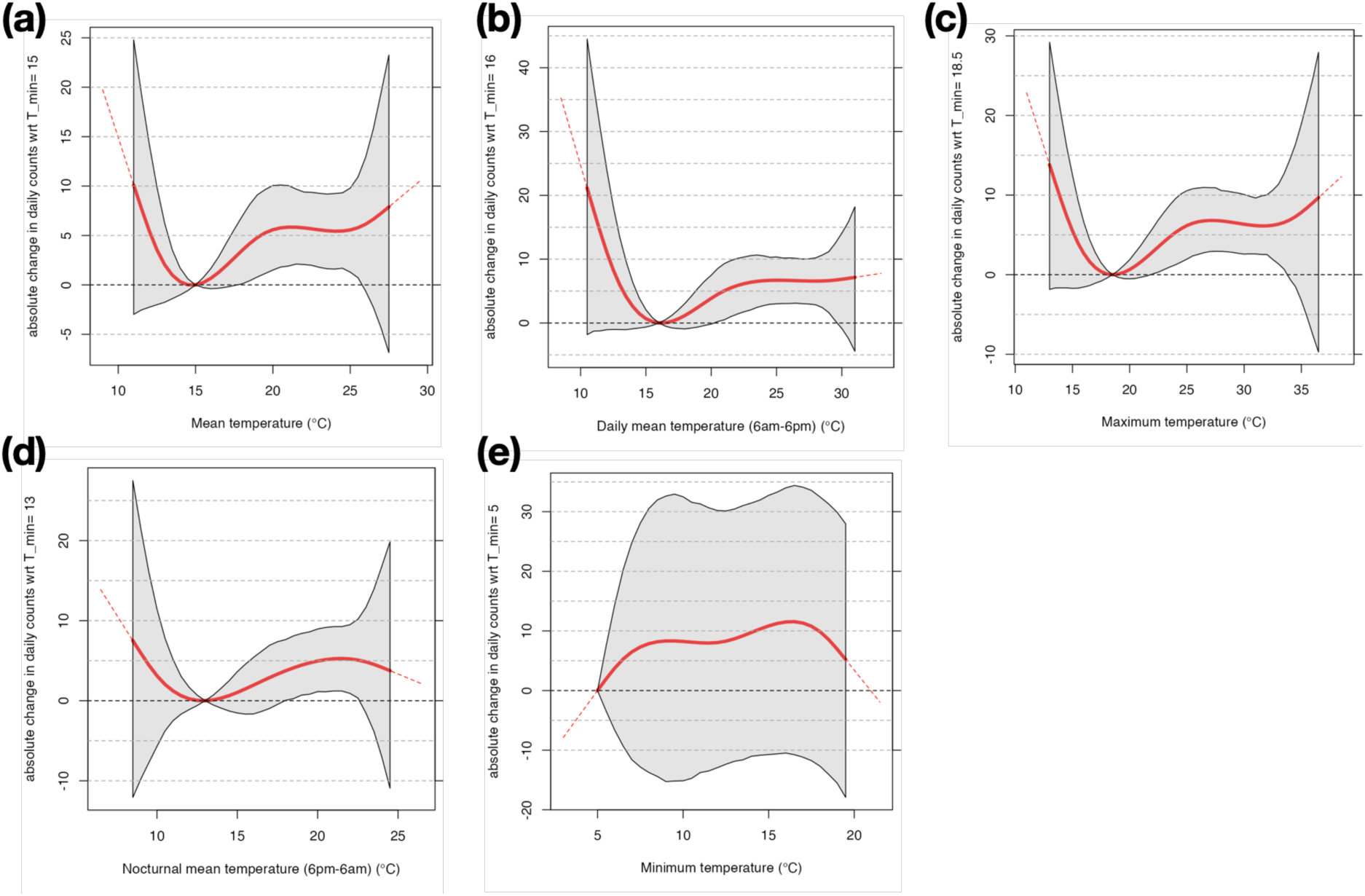
Cumulative exposure-response function. obtained using different temperature indicators: **(a)** mean temperature (as in the main analysis); **(b)** mean temperature during the day (6am - 6pm); **(c)** maximum temperature; **(d)** nocturnal mean temperature (6pm - 6am); **(e)** minimum temperature.

### Gaussian and quasi-Poisson regression

We fitted the regression model assuming a Gaussian distribution, which is the most common choice in forecasting models. Studies focusing on estimating the association with temperature (however, without making forecasts), usually adopt a quasi-Poisson distribution^15^. We found that the shape of the exposure-response function estimated from our dataset was not sensitive to the choice of the distribution (Gaussian or quasi-Poisson, Fig. S6); besides, our time-series included large daily counts (more than 100). For these reasons, we decided to use a Gaussian distribution. However, we acknowledge that this may not be a suitable choice when dealing with a time-series with very low case numbers.

**Fig. S6.**
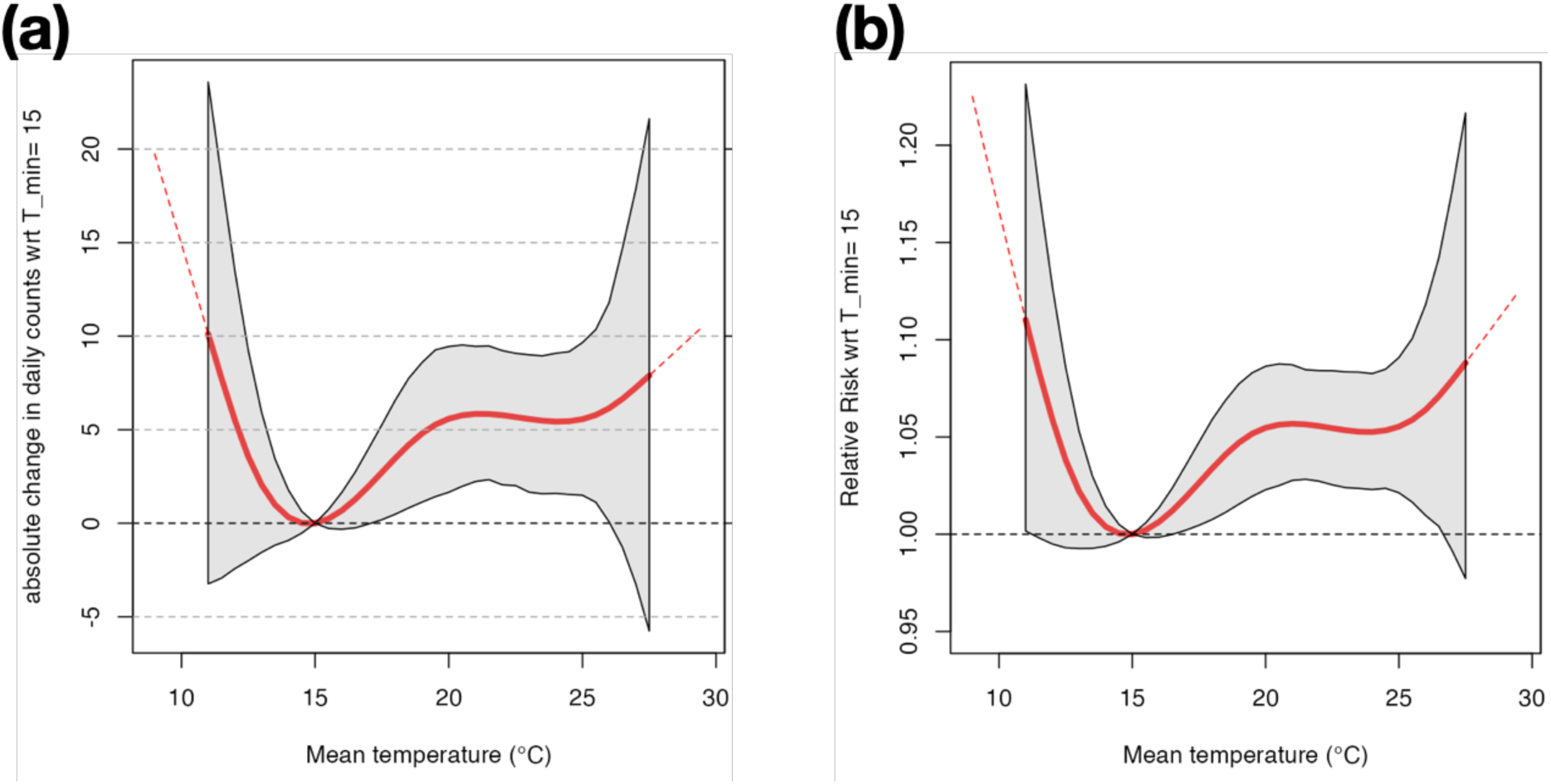
Cumulative exposure-response function. obtained using different distribution families. **(a)** Model assuming a Gaussian distribution (as in the main analysis) **(b)** Model assuming a quasi-Poisson distribution.

### Observed vs forecasted temperature data

We had access to forecasted temperature data for summer 2022. These forecasts were generated through the COSMO model at MeteoSwiss, providing forecasts up to 6 days ahead. For each forecast reference time (100 days, from 1st of June 2022 to 31st of August 2022), we considered temperature quantiles (10th, 20th, 50th, 80th & 90th) for each lead-time from +0 days to +5 days, for the air temperature at 2m above ground at the Zollikofen station. Post-processing on the gridded ensemble forecast was done as follows: we interpolated the temperature grid to only the Zollikofen measurement station; we computed the quantiles from the 21 ensemble members; we took the daily means (as the original data is hourly). A sample of the data for the first two weeks in June 2022 is illustrated in Fig. S7. The quantile intervals generally correspond to variations of 2 °C compared to the median value.

**Fig. S7.**
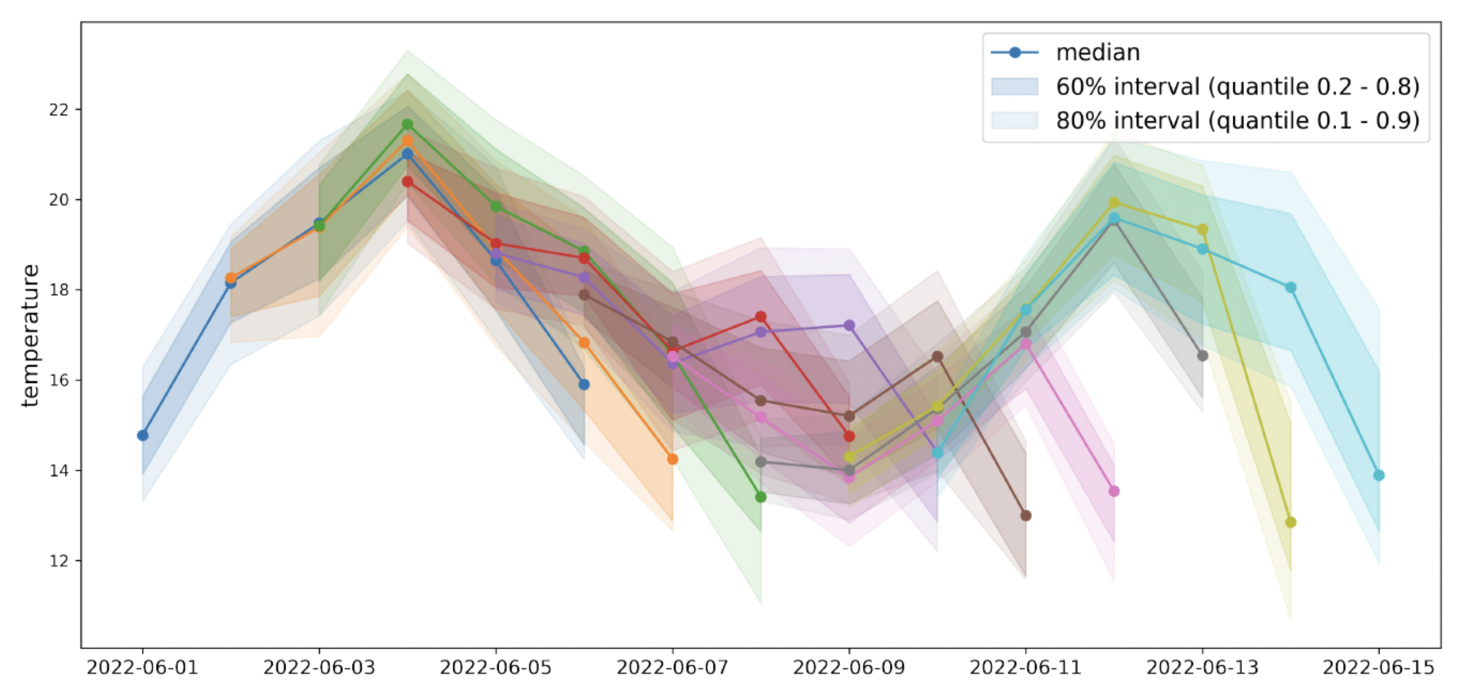
Forecasted temperature data. Lines indicate median values, shaded areas indicate quantiles (20th - 80th and 10th - 90th). Each color refers to a given reference time, and contains 6 lead times (from +0 days to +5 days).

First, we compared the forecasted temperature data with the observed temperature data used in our main analyses, registered from the Zollikofen station. We found that the median forecasted temperature data are aligned quite well with the observed temperature data (Fig. S8). The best alignment is found for a lead time of 0 days, but good agreement is also found up to a lead time of 4 days. For lead time equal to 5 days, the forecasted temperature data underestimate the observed temperature data.

**Fig. S8.**
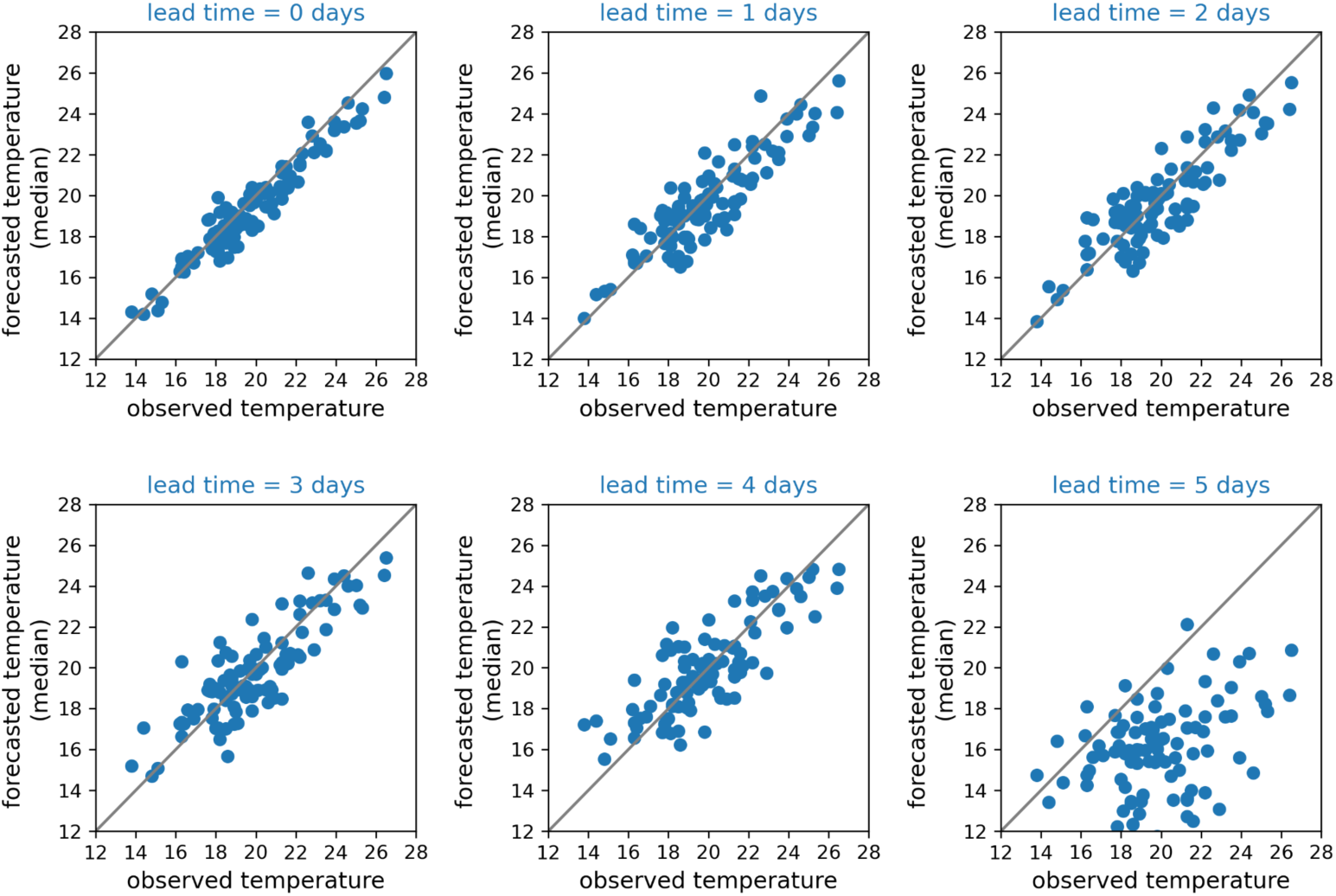
Comparison between (median) forecasted temperature data and observed temperature data. For each panel, we show a scatter plot with the observed temperature (x-axis) and the corresponding forecasted temperature (y-axis) obtained with a given lead time (from 0 to 5 days in advance).

Secondly, we generated hospital forecasts using as input the median forecasted temperature, compared to using observed temperature. To quantify the impact of the uncertainty in the forecasted temperature, we also generated hospital forecasts using the upper or lower quantiles of the forecasted temperature. As expected, the number of hospital visits are generally slightly higher when using the upper bound of the forecasted temperature instead of median values (Fig. S9). However the upper range of the prediction interval does not increase largely. In other words, the prediction interval for the hospital visits obtained with the median temperature already captures most of the uncertainty, and including uncertainty in the temperature values in input does not lead to a significant extension of the prediction interval.

**Fig. S9.**
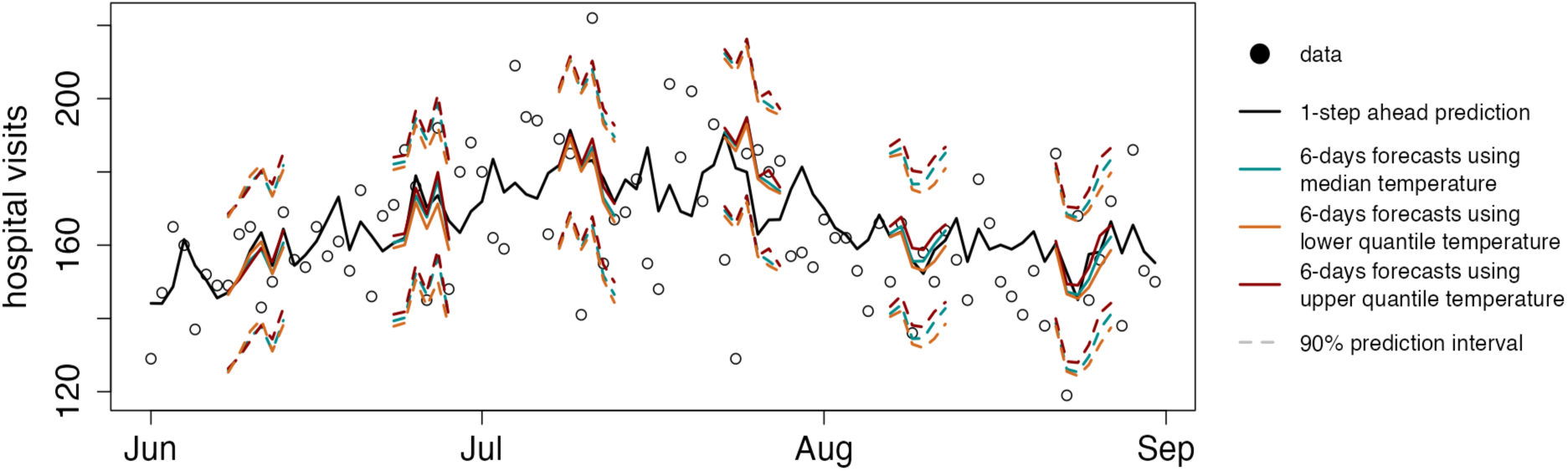
Hospital forecasts using observed vs forecasted temperature data as input. **Dots represent observed data of emergency room visits (ERVs). Black line indicates the 1-step ahead prediction for reference. Colored lines indicate** 6-days forecasts obtained in six different time windows, obtained using as predictor the median forecasted temperature (green), the 10% quantile (orange) or the 90% quantile (red). Continuous line indicate the point-prediction, dashed lines indicate the 90% prediction interval.

We computed the RMSE of the point predictions generated by three models (using median, upper or lower quantiles of the forecasted temperature as input), and compared them with the original model using observed temperature data. We found that the average RMSE over 6-days rolling forecast windows remained quite similar across model inputs (Fig S10).

**Fig. S10.**
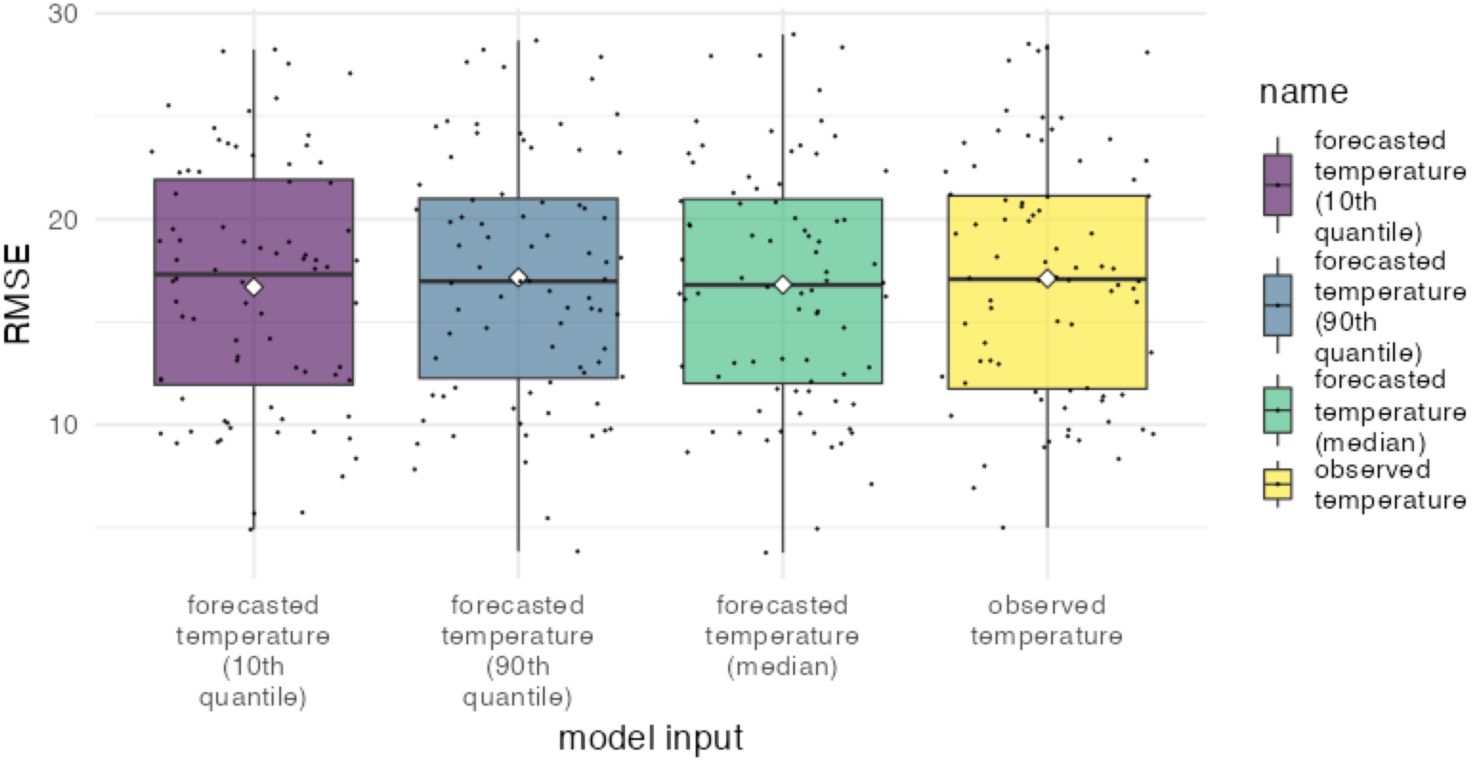
Forecast errors using observed or forecasted temperature as input. Root-mean-squared-error of hospital forecasts against observed hospital data, evaluated in time windows with a length of 6 days, obtained with a model using 10% quantile, 90% quantile or median of forecasted temperature, or observed temperature.

## Notes

### Competing Interest Statement

The authors have declared no competing interest.

